# Real-time, multi-pathogen wastewater genomic surveillance with Freyja 2

**DOI:** 10.1101/2025.07.26.25332245

**Authors:** Joshua I. Levy, Praneeth Gangavarapu, Dylan A. Pilz, Maryam Ahmadi Jesvaghane, Allison Steedman, Mark Zeller, Karthik Gangavarapu, Daniel McDonald, Kevin Libuit, Curtis Kapsak, Erin L. Young, Shelesh Agrawal, Laura Orschler, Martin Hölzer, Elyse Stachler, Daniel J. Park, Angie S. Hinrichs, Louise C. Laurent, Rob Knight, Kristian G. Andersen

## Abstract

Case-based infectious disease surveillance is fundamental to public health, but is resource-intensive, logistically complex, and prone to sampling bias. Wastewater testing and sequencing have increasingly been used for population-scale monitoring of pathogen dynamics, including in low-resource settings. Broader adoption of wastewater genomic surveillance, however, is limited by a lack of flexibility across sequencing platforms and approaches, and adaptability to additional pathogens. Here, we describe “Freyja 2”, an integrated bioinformatics tool enabling robust real-time inference of pathogen lineage prevalence and growth dynamics from wastewater and other complex samples. In Freyja 2, we develop new methods for estimating lineage prevalence and growth rates, and demonstrate robustness across common sequencing platforms and to low genomic coverage. By incorporating global pathogen data streams, we extend Freyja 2 to support multi-pathogen surveillance. We demonstrate tracking of multiple recent or ongoing public health emergencies, including COVID-19, mpox, and H5N1 influenza, revealing unreported diversity and lineage co-circulation.

## Introduction

Emerging, re-emerging, and endemic infectious diseases continue to threaten communities globally^1,2^. Outbreak tracking and response mostly rely on resource-intensive, clinical case-based surveillance that can be subject to structural biases^3,4^. Consequently, timely and actionable testing and sequencing at scale remain challenging or cost-prohibitive in many settings^5,6^. Strengthening and expanding our public health monitoring systems will require developing passive surveillance technologies that are community representative, low-cost, and adaptable to diverse pathogens^7–9^.

Wastewater transport systems are among our most powerful and cost-effective sources to understand infectious disease burden and obtain other critical public health information^10–12^. Many pathogens, including SARS-CoV-2, Zika virus, influenza viruses, and mpox virus (MPXV) are shed via stool and/or urine from infected individuals and are detectable downstream in wastewater collections^13^. These samples can be used to detect and quantify the community burden of multiple co-circulating pathogens passively, with minimal resource requirements^14^. Sequencing wastewater samples enables genomic characterization of pathogen strains, variants, and lineages in the community and reveals evolutionary and growth trends, at similar cost to sequencing a single clinical isolate^11,15–17^. Genomic analysis of wastewater and other complex samples is challenging, however, because they contain mixtures of co-circulating pathogens and their variants^18,19^. Further, nucleic acids from these sources are typically sparse and highly fragmented, often resulting in low sequencing coverage and depth. To fully capture pathogen diversity of community biospecimens and accurately characterize sample composition from mixed and other complex samples there is therefore a need for new, flexible, and scalable approaches^20–23^.

To enable analysis of SARS-CoV-2 lineage dynamics from wastewater, we previously developed “Freyja”, a bioinformatic tool for estimating lineage prevalence from complex samples^24^. We found that Freyja robustly tracked SARS-CoV-2 lineage dynamics, enabled earlier detection of variants of concern, and identified cryptic variant circulation^24^. Using both real and simulated data, Freyja has been shown to outperform other methods in estimating virus lineage and variant prevalence^25–28^. Since its initial release, Freyja has been widely adopted by the public health community, from local health departments to national and international surveillance programs. Across numerous longitudinal wastewater sequencing efforts, Freyja has enabled earlier detection of emerging variants compared to case-based surveillance and expanded tracking of evolutionary dynamics, including in low- and middle-income countries^24,25,29–32^.

Here we introduce Freyja 2, a robust and scalable toolkit for real-time genomic and epidemiological analysis that enables multi-pathogen surveillance from wastewater and other complex samples. We establish automated workflows that incorporate up-to-date information on pathogen diversity and support analyses across common computing platforms, with minimal requirements for bioinformatics expertise and computational resources. We develop new inference methods for estimating lineage prevalence that improve accuracy and robustness, including for incomplete and low-quality data across common sequencing platforms, and demonstrate a new approach to infer lineage-specific growth rates from longitudinal wastewater data. Finally, we extend Freyja into a real-time multi-pathogen framework to gain insights into several public health emergencies, including the ongoing multi-country mpox outbreak and influenza A/H5N1 epizootic. Taken together, these advances enable robust multi-pathogen genomic surveillance and strengthen community-level pathogen monitoring.

## Results

### Robust lineage prevalence inference from complex samples underpins global wastewater-based pathogen monitoring

To infer lineage prevalence from complex samples containing highly similar and fragmented pathogen genomes observed in wastewater and other mixed samples, Freyja exploits the relationship between single nucleotide polymorphism (SNP) frequency and the overall frequency of pathogen lineages defined by those SNPs, as in the original Freyja method (Freyja 1):

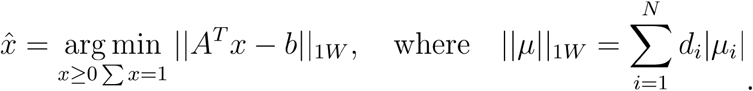

To recover lineage prevalence, *x*, Freyja encodes the sequence of SNPs that define each lineage into binarized “lineage barcodes”, *A*, and observed SNP frequencies into a vector *b* ^24^. This constrained optimization method uses a weighted L1 norm, denoted as 1*W*, with site-specific sequencing depth, *d*_*i*_, to solve for the lineage proportions that best explain the empirical SNP frequency profile, while ensuring prevalence estimates are nonnegative and sum to 1. This approach allows for flexible and accurate inference of pathogen prevalence from complex samples and has substantial robustness to incomplete genome coverage.

### Automated workflows enable real-time, reproducible analyses across computing platforms

Freyja has been widely deployed for wastewater genomic surveillance of SARS-CoV-2^29,33–36^. To stay current with the continued evolution of SARS-CoV-2 and corresponding designation of new virus lineages through efforts such as PANGO^37^, we established automatic daily updates using the latest global phylogenetic tree via UShER^38,39^ and integration of updated variant definitions into lineage barcodes in Freyja 2 (**Fig. 1a**). These barcodes are time-stamped, versioned, and stored in a dedicated Freyja-data repository (**Fig. 1b**)^40^. Updates occur client-side within seconds, ensuring that Freyja analyses always incorporate up-to-date SARS-CoV-2 variant and lineage classifications. Barcode version information is easily checked, and corresponding versioned barcodes from Freyja-data can be supplied for backwards compatibility and reproducibility.

**Figure 1.**
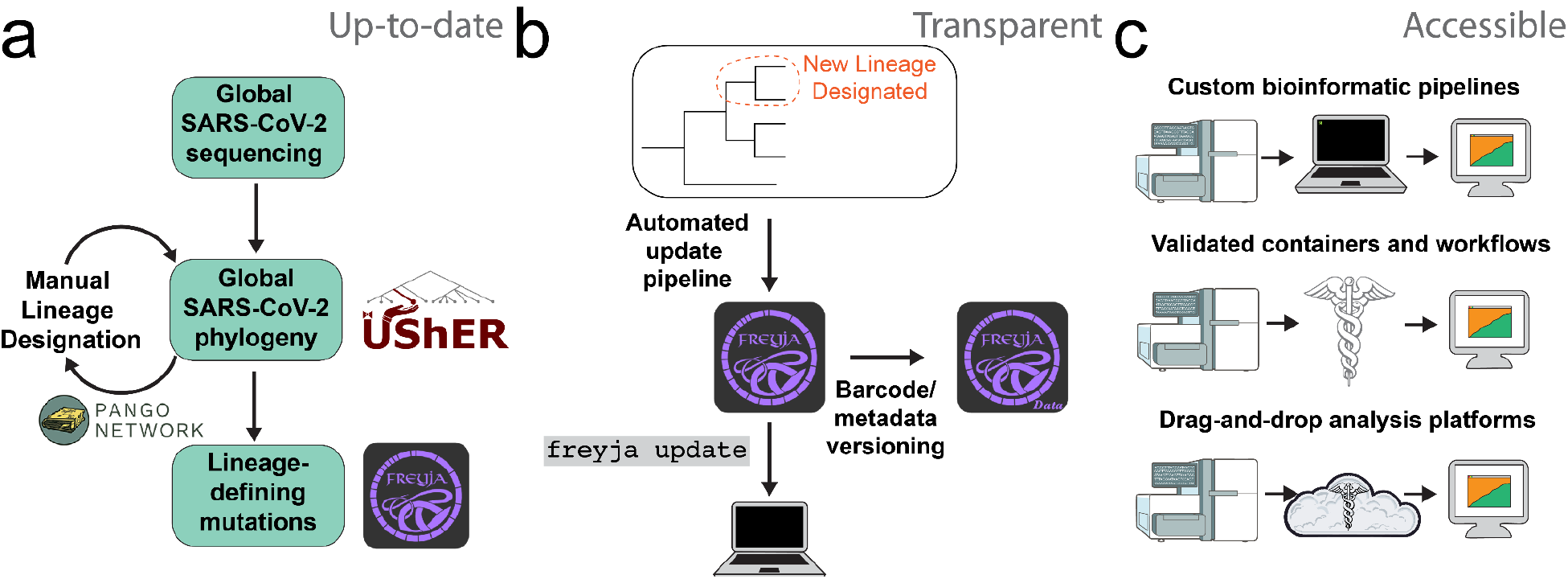
Community resources enable SARS-CoV-2 wastewater surveillance at scale. **a**. As new sequences become available, they are placed onto the global phylogeny and assigned to an existing clade. Novel lineages are manually designated by the Pango Network and associated taxa are re-assigned in the global phylogeny, maintained by the UShER team. Mutation paths associated with each lineage root are then extracted and used for lineage-barcoding. **b**. Upon designation of a novel lineage, an automated pipeline is used to update the lineage barcode “library”. This new “library” is then immediately available to users via a simple update command, and with versioning of all barcodes and variant grouping metadata. **c**. Freyja is available via pre-built, web interface-enabled tools, and is integrated within public health wastewater bioinformatic workflows.

To support broad accessibility regardless of end-user bioinformatics experience, we developed multiple options for end-to-end processing of wastewater sequencing data, including automated generation of report-ready visualizations of lineage prevalence trends (**Fig. 1c**; **Supp. Table 1**). For users with bioinformatics training, Freyja can be installed via open-source package channels such as bioconda and run using custom command-line scripts and pipelines. For public health contexts requiring complete pre-configured and validated workflows, we established collaborations with organizations including the State Public Health Bioinformatics (StaPH-B) workgroup (>170,000 downloads of the StaPH-B Docker container as of May 2025)^41^ to make validated workflows freely available. Freyja has also been independently integrated into major analysis pipelines including Aquascope, Cecret, and poreCov^27,42–44^. Because many public health laboratories operate without in-house bioinformatics support, we also packaged these workflows for user-friendly deployment through graphical, point-and-click interfaces such as Terra and Galaxy^45,46^. These core resources and validated workflows enable broad-scale implementation of environmental genomic surveillance, supporting initiatives such as the CDC National Wastewater Surveillance System (NWSS) in the United States^47^ and the National Institute for Communicable Diseases of South Africa national wastewater program^29^.

### Explicit error modeling and coverage accounting improve lineage prevalence inference accuracy

The quality and genome coverage of sequencing from wastewater and other complex mixtures can vary significantly, introducing platform-specific errors, which limit the accuracy of pathogen lineage prevalence inference. To explicitly account for error rate differences across sequencing platforms and lineage identifiability as a result of partial genome coverage, we developed a new regularized method in Freyja 2 that adjusts to empirical error profiles:

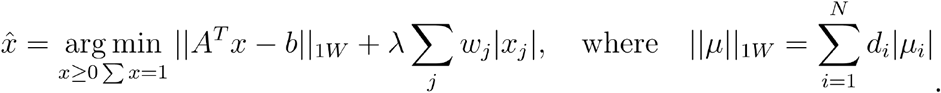

This method uses a penalty, *λ*, to explicitly balance the prevalence attributed to each lineage with the residual between the inferred sample representation and empirical data. Using a lineage-specific adaptive weighting, *w*_*j*_, (**Fig. 2a**, orange box) ensures consistent variable selection (i.e., the solution converges to the true solution given sufficient observations^48^) and enables detection of low frequency pathogen variants while limiting false detections. Second, to ensure Freyja 2 analyses reflect variation in lineage identifiability due to coverage differences and therefore accurately represent sequenced mixtures, we mask SNPs in the barcode matrix that occur at genomic positions without sequencing coverage (**Fig. 2d**),

**Figure 2.**
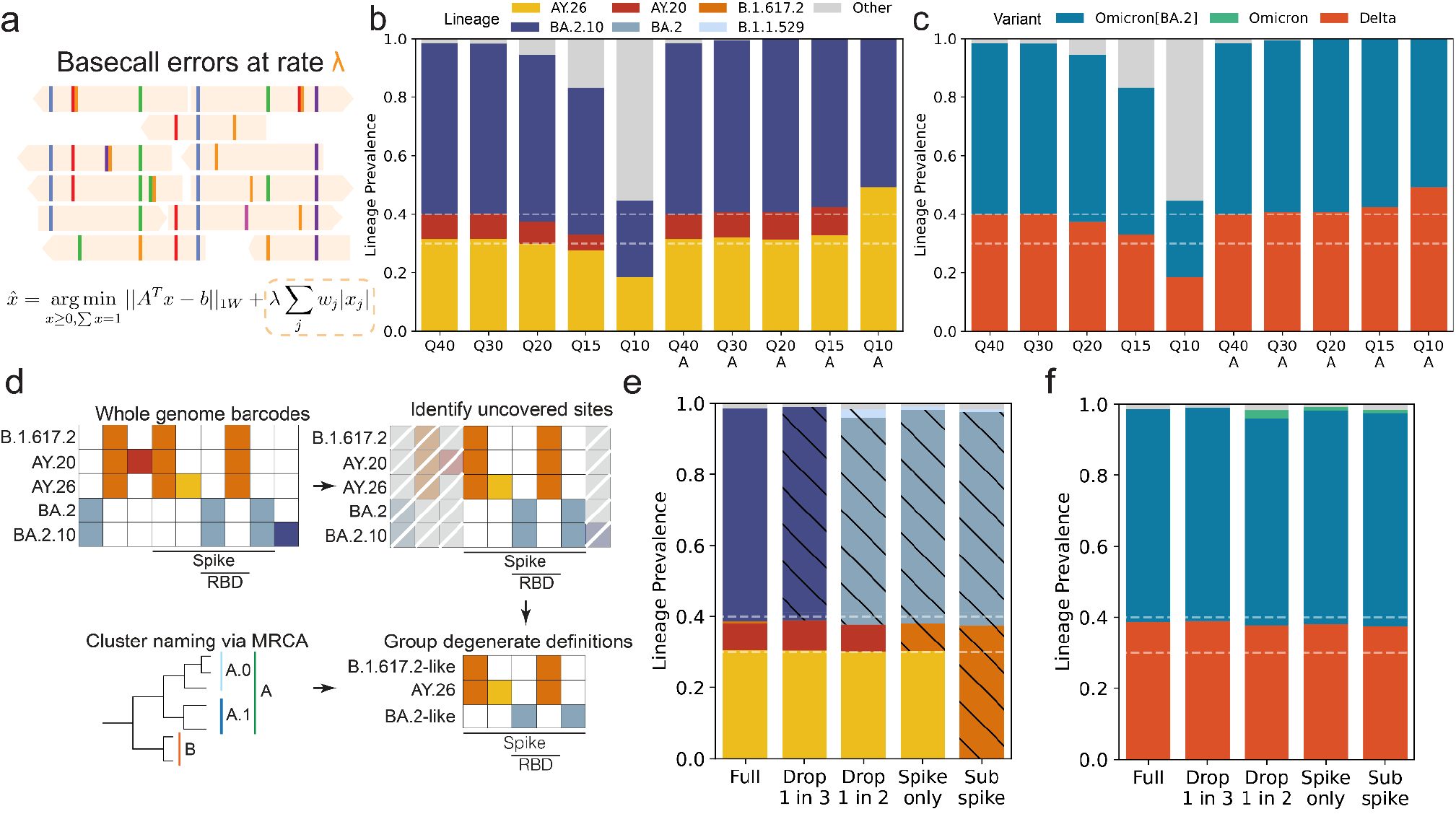
Robust lineage prevalence inference by explicitly incorporating genomic coverage and sequencing error rates. **a**. Base-calling errors can lead to incorrect identification of virus diversity, but are accounted for in Freyja 2 using an adaptive, regularized model, while still following nonnegativity and unit sum constraints. **b-c**. Example results for a simulated 10/30/60 mixture sample, for a range of sequencing qualities. **b**. Lineage and **c**. variant level deconvolution results, with adaptive regularization (Freyja 2) results denoted by an “A” under the simulated sequencing quality. **d**. Schematic showing how genomic coverage can make some lineages indistinguishable, provided available data. Freyja 2 automatically accounts for genomic coverage when naming lineage groups. **e-f**. Example Freyja 2 results for the simulated 10/30/60 mixture sample using a range of possible sequencing coverages, from full genome coverage, to partial amplicon dropout, to spike-only coverage. Black hatching indicates that the identified lineage corresponds to a class of lineages with the named lineage as its most recent common ancestor. **e**. Lineage-specific and **f**. variant-summarized deconvolutions for the considered sequencing schemes.

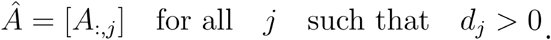

If missing sites cause some lineages to be identical (i.e. 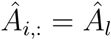) with respect to the remaining SNPs, they are merged and named in accordance with their most recent common ancestor (MRCA).

To validate the effectiveness of our regularized method in controlling for sequencing errors, we simulated amplicon sequencing of mixed lineage samples (**Supp. Fig. 1**) using error rates spanning commonly used platforms, varying base qualities from Q40 to Q10. Next, we determined inference accuracy at the resolution of individual lineages (e.g. SARS-CoV-2 lineage B.1.1 or BA.2.10) and of variants, which include all lineages with a shared parent (e.g. BA.2.10 descends from the now de-escalated Omicron BA.2 variant of concern), and compared the results of Freyja 2 relative to Freyja 1. For Q20 (comparable to raw sequencing data from current Oxford Nanopore flow cells) and lower quality sequencing, we found that Freyja 2 led to decreases in lineage-level inference errors of more than 50% (**Fig. 2b, Supp. Fig. 2a**; Q20: from 0.05 to 0.02; Q15: 0.17 to 0.03; Q10: 0.44 to 0.20). We observed even more pronounced decreases in the number of detections and overall prevalence attributed to false positive, “off-target” lineages (Q20: Off target prevalence from 0.048 to 0.003; Q15: 0.16 to 0.01; Q10: 0.45 to 0.04; **Supp. Fig. 2c,d**), with only small decreases in lineage-level sensitivity (i.e., probability of true detection of a lineage; **Supp. Fig. 2b**). All false negatives occurred as lineage frequency approached the sequencing error rate (**Supp. Fig. 3**). At the variant-level, similar results were observed for Q20 and lower quality (**Fig. 2c, Supp. Fig. 2a-c**). We observed reduced estimate error (Q20: 0.02 to 0.006, Q15: 0.05 to 0.01, Q10: 0.17 to 0.11), minimal impact on sensitivity, and a near complete reduction of off-target hits and prevalence, even for Q10 reads.

**Figure 3.**
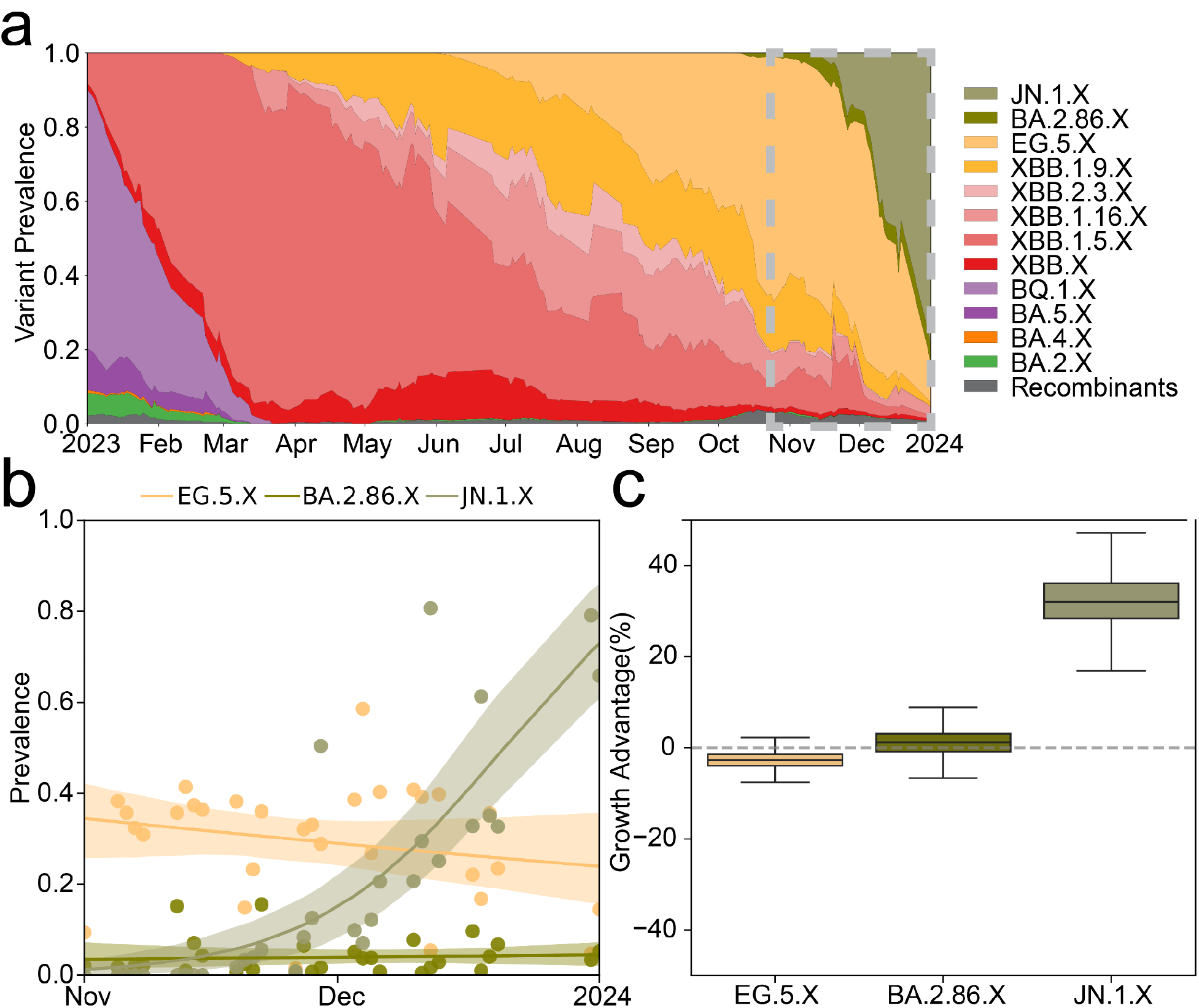
Tracking and quantifying lineage dynamics from wastewater. **a**. Longitudinal lineage prevalence during 2023 at the Point Loma wastewater treatment plant in San Diego. Dotted grey rectangle indicates the region over which growth advantage is calculated. **b**. Raw lineage prevalence estimates and inferred logistic model for each lineage, with bootstrap prediction intervals shaded. **c**. Distribution of bootstrap estimates of the growth advantage associated with each lineage.

To validate the utility of using available genomic coverage to characterize lineage identifiability during lineage prevalence inference, we simulated mixtures replicating a range of amplicon-based sequencing scenarios, from whole genome to sub-spike schemes, and amplicon dropout (**Fig. 2d**). As intended, Freyja 2 returned lineage groupings when lineage distinguishing mutations of the genome were not available (e.g., BA.2.10 merged with BA.2 and other BA.2 descendants and named BA.2-like after the group’s MRCA), and aggregate grouping prevalence reflected the sum of constituent lineages’ prevalence (**Fig. 2e**). In random amplicon dropout simulations, we found that accounting for lineage identifiability consistently reduced lineage-level inference error, and provided accurate estimates at as high as 40% amplicon dropout (mean error of 0.04 at 40% dropout; **Supp. Fig. 2e**). We observed decreased off-target (including ancestral lineage) detections and lineage prevalence (<1 on average at 25% dropout; **Supp. Fig. 2f,g**), but as lineages were grouped due to lack of genome coverage, we observed an expected decrease in lineage-level sensitivity (**Supp. Fig. 2h**), since not accounting for identifiability can lead to multiple lineage detections when they are equally likely based on the data, even if they are indistinguishable given available genome coverage. At the variant-level, the new method almost completely eliminated estimate error, maintained near-perfect sensitivity, and had low off-target variant prevalence, including for sub-spike data (**Fig. 2g, Supp. Fig. 2g**). Even for as much as 70% amplicon dropout, variant-level estimates were highly accurate, with very few false negatives or false positives.

### Freyja 2 enables direct inference of lineage-specific growth rates from wastewater

A critical limitation of current wastewater genomic surveillance tools, including Freyja 1, is that they can not quantify increases in lineage fitness, which are typically characterized using clinical sequence data and may stem from diverse factors including changes in transmissibility or host immune evasion. To quantify lineage-specific growth advantage directly from longitudinal genomic surveillance of wastewater and other complex samples (**Fig. 3a**), we developed an approach to compute the lineage selection coefficient, *s*, which relates the effective reproduction number *R*^*^ of a lineage of interest to the effective reproduction *R*^*B*^ number of one or more co-circulating lineages

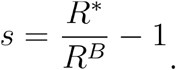

Our approach models the exponential growth dynamics of lineage transmission, is invariant to virus shedding, and reduces to a logistic model. To validate this approach, we simulated variant emergence dynamics using a two-lineage model including temporal virus shedding following infection and a binomial process to simulate observation of reads belonging to the lineage of interest (**Supp. Fig. 4a,b**). We found that model estimates became more accurate as the number of reads at sites distinguishing the two lineages increased. Although increased sampling frequency to more than once per week decreased variance in the estimated growth rate advantage, sampling more than twice per week was of limited utility, including for estimates using only the early stage of the outbreak (**Supp. Fig. 4c,d**), suggesting that sampling frequencies compatible with established wastewater utility capabilities and environmental sampling programs will be sufficient. To validate this approach on longitudinal genomic surveillance, we analyzed wastewater sequencing data from San Diego^49^ (**Fig. 3a**). We found that lineage prevalence followed logistic-like trajectories (**Fig. 3b**), and using a bootstrap resampling approach, we identified a significant growth advantage for JN.1.X during this time period (**Fig. 3c**). Freyja 2 estimated a growth advantage of 31% (Bootstrap 95% CI: [23%,46%]) during this time period in San Diego, closely matching reported clinical sequence-based estimates, including for France, the United Kingdom, and Spain^50^.

### Development of a generalized framework enables multi-pathogen sequencing analyses

When we initially developed Freyja, we focused on SARS-CoV-2 given the rapid emergence and co-circulation of multiple variants of concern^24,51^; however, many other key pathogens and their variants can be detected in wastewater samples^13,52,53^. To generalize the usage of Freyja 2 for other pathogens, we established a streamlined computational framework for building pathogen-specific lineage barcodes (**Fig. 4**). For any pathogen of public health interest, such as MPXV, Zika virus, or avian influenza A/H5N1 virus (**Fig. 4a**), the framework leverages existing community resources to determine lineage defining mutations that can be directly used within Freyja 2. To create lineage barcodes for each pathogen, we first ingest lineage annotations (**Fig. 4b**) and annotate mutations onto phylogenetic trees (**Fig. 4c**). For pathogens with existing lineage designations and associated nomenclature, including for MPXV^54^, we integrate community genomic epidemiology resources including Nextstrain, PANGO^55^, and GISAID^56^ (**Supp. Fig. 5a-c**). For pathogens with limited or no consensus cladistic nomenclature, including Zika virus and the influenza A/H5N1 cattle epizootic, we perform automated lineage assignment with autolin^55^ and incorporate manual lineage designations when available (**Supp. Fig. 5b,c**). The resulting mutation-annotated trees are then converted to lineage barcodes using Freyja (**Fig. 4d**).

**Figure 4.**
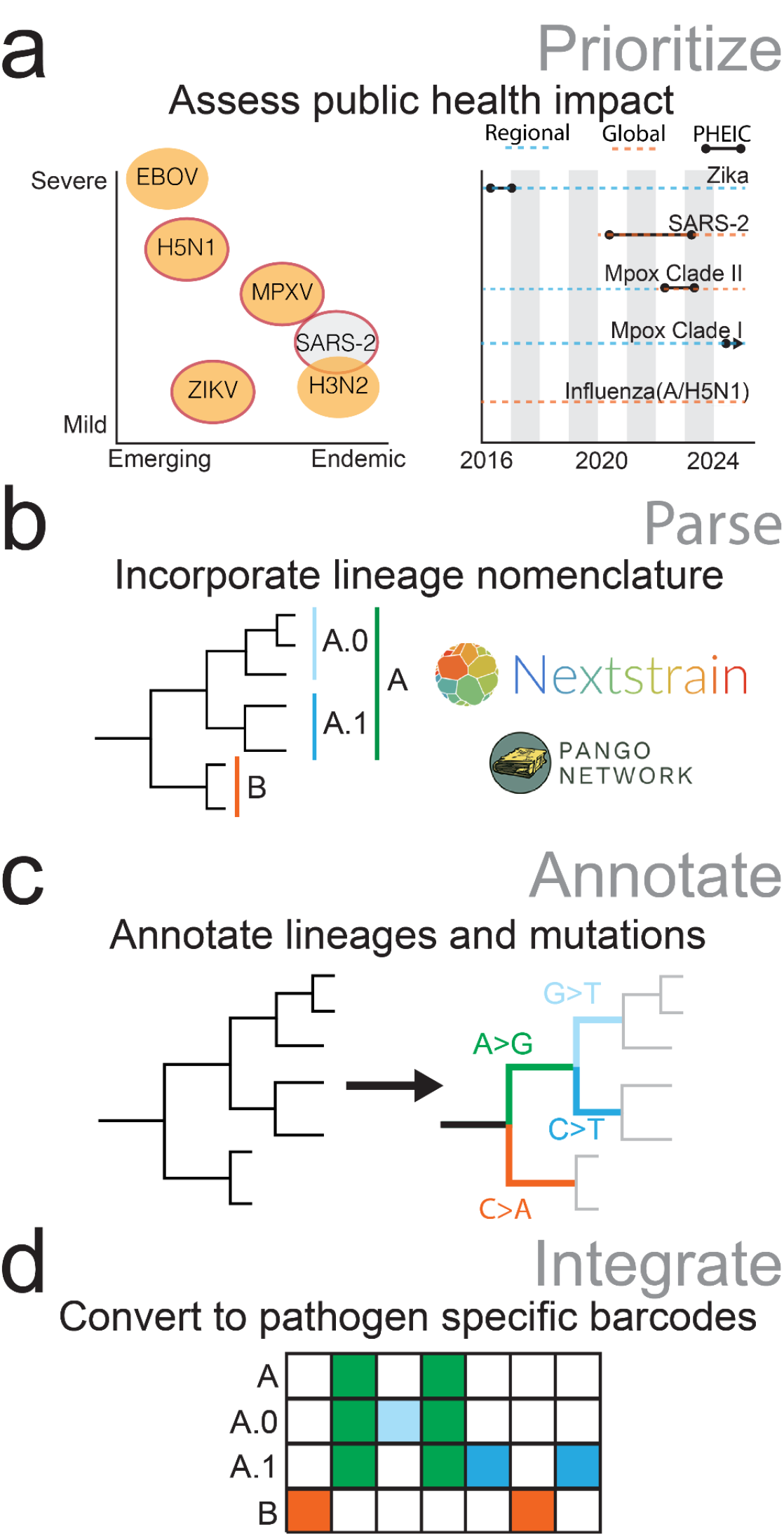
Extending Freyja for multi-pathogen wastewater genomic surveillance. **a**. Identifying key pathogens impacting public health that are detectable in wastewater, with red outlines for pathogens studied in this work (left). Recent public health emergencies of international concern (PHEICs) and outbreaks with human pandemic potential (right). **b**. Parsing community nomenclature and associated resources enables real-time monitoring of target pathogens. **c**.,**d**. The BarcodeForge workflow performs **c**. annotation of mutations onto the phylogeny to determine the path leading to each lineage and **d**. processing of mutation paths into lineage-defining barcodes.

To validate our framework, we performed *in silico* validation for each pathogen (**Supp. Fig. 6**). Using high-quality representative sequences for each lineage, we simulated amplicon sequencing of lineage mixtures, identified cases of amplicon failure due to primer mismatches, and after analysis with Freyja 2, determined inference accuracy. We then further validated with real sequencing data of two-component spike-in mixtures of MPXV and Zika virus, including multiple common sequencing platforms ranging in read quality (**Supp. Fig. 7**). For MPXV, we found that Freyja 2 enabled robust and accurate estimation of lineage prevalence, with an average lineage prevalence error of 0.006, no false positives or false negatives across all tested mixtures, and reliable detection of lineages even as low as 1% prevalence (**Supp. Fig. 7a,b**). For Zika virus, we observed negligible differences in estimated prevalence across high (Q30-Q40) and low quality (Q15) amplicon-based sequencing (**Supp. Fig. 7c,d**). Freyja 2 provided accurate results for all samples regardless of the choice of sequencing platform, with no false positives or negatives. These findings demonstrated that Freyja 2 can be readily generalized to other pathogens, while maintaining the accuracy and robustness for lineage and clade prevalence inference, as originally demonstrated for SARS-CoV-2. Each of these barcodes, and others including for measles and RSV, are publicly available (github.com/andersen-lab/Freyja-barcodes). The barcode generation workflow, BarcodeForge (github.com/andersen-lab/BarcodeForge), is freely available for community use.

### Insights into multi-pathogen outbreak dynamics using sequencing of complex samples

To assess Freyja 2’s utility for real world multi-pathogen genomic surveillance, we analyzed aggregate collections from two public health emergencies: the ongoing multi-country mpox outbreak^57^ and the influenza A/H5N1 epizootic in cattle in the United States^58^. To study circulating MPXV lineage prevalence using wastewater, we obtained and analyzed publicly available wastewater sequencing data from Germany, including samples from May–August 2022^59^. During this period, Germany had the most clinical MPXV clade IIb lineage B.1 detections in Europe, but had no detections of its parent, lineage A (**Fig. 5a**), despite Germany having among the earliest known mpox cases on the continent (**Supp. Fig. 8a**) and a sequencing rate of over 20%^60–62^. Among the wastewater sequencing data we analyzed, which was generated using amplicon-based approach on the Ion Torrent platform, we found all samples were constituted of a single lineage grouping of Clade IIb, initially A-like and then B.1-like, matching clinical trends observed elsewhere in Europe during this time period (**Fig. 5b**)^60^. As many of these samples had low genomic coverage, Freyja 2 identified groups of lineages that were not distinguishable, with more specific groupings as coverage increased (**Fig. 5c**), as enabled by Freyja 2’s adaptive lineage-merging strategy. Notably, Freyja 2 yielded stable lineage groupings even for samples with as little as 10% genome coverage, highlighting its utility in complex environmental samples where dropout and degradation are common. Due to limited genome coverage, the early lineage A-like detection could not be distinguished from many of its descendants. These results were consistent with lineage assignments from an earlier analysis of the same dataset using consensus genome sequence construction and phylogenetic placement, where consensus sequences for samples collected in August 2022 clustered within the B.1 lineage in the original study^59^. The ability of Freyja 2 to infer consistent lineage groupings across diverse platforms and amplicon schemes demonstrates its robustness to differences in available coverage and sequencing quality.

To track the ongoing A/H5N1 epizootic, which has already led to infections in hundreds of herds (**Fig. 5d**) but has caused only sporadic human cases, wastewater collections are poorly suited for tracking virus evolution. However, during A/H5N1 infection cows shed the virus via milk, and these viruses can be detected and sequenced, similarly to wastewater collections^63^. We used publicly available sequencing of milk to monitor infections at a range of scales, from individual cows, to bulk tanks from individual farms, to retail milk with contributions from multiple individual farms (**Supplementary Data**). Using a combination of existing hemagglutinin clade designations^64^ and new lineages in the B3.13 cattle epizootic clade^58^ (**Supp. Fig. 5c**), we found that although the majority of milk samples were composed of only one lineage, we detected two or more lineages in about 10% of samples (**Fig. 5e, Supp. Fig. 8b**). In pooled milk samples including bulk (generally from a single farm) and retail milk (generally from multiple farms), the rate of two or more lineage detections was higher than that of samples labeled as “milk”, with nearly 15% of bulk samples, and 40% of retail milk samples containing more than one lineage. Among samples listed as “milk”, assumed to be derived from individual cows^58,65,66^, we did still identify multiple lineages in more than 10% of samples, suggesting some herds may have had multiple simultaneous outbreaks (**Fig. 5f**). These findings indicate that Freyja 2 uncovered previously unexplored diversity in complex samples of both MPXV and A/H5N1, and that leveraging composite agricultural collections in addition to wastewater can enable characterization of human and zoonotic pathogen diversity and circulation.

**Figure 5.**
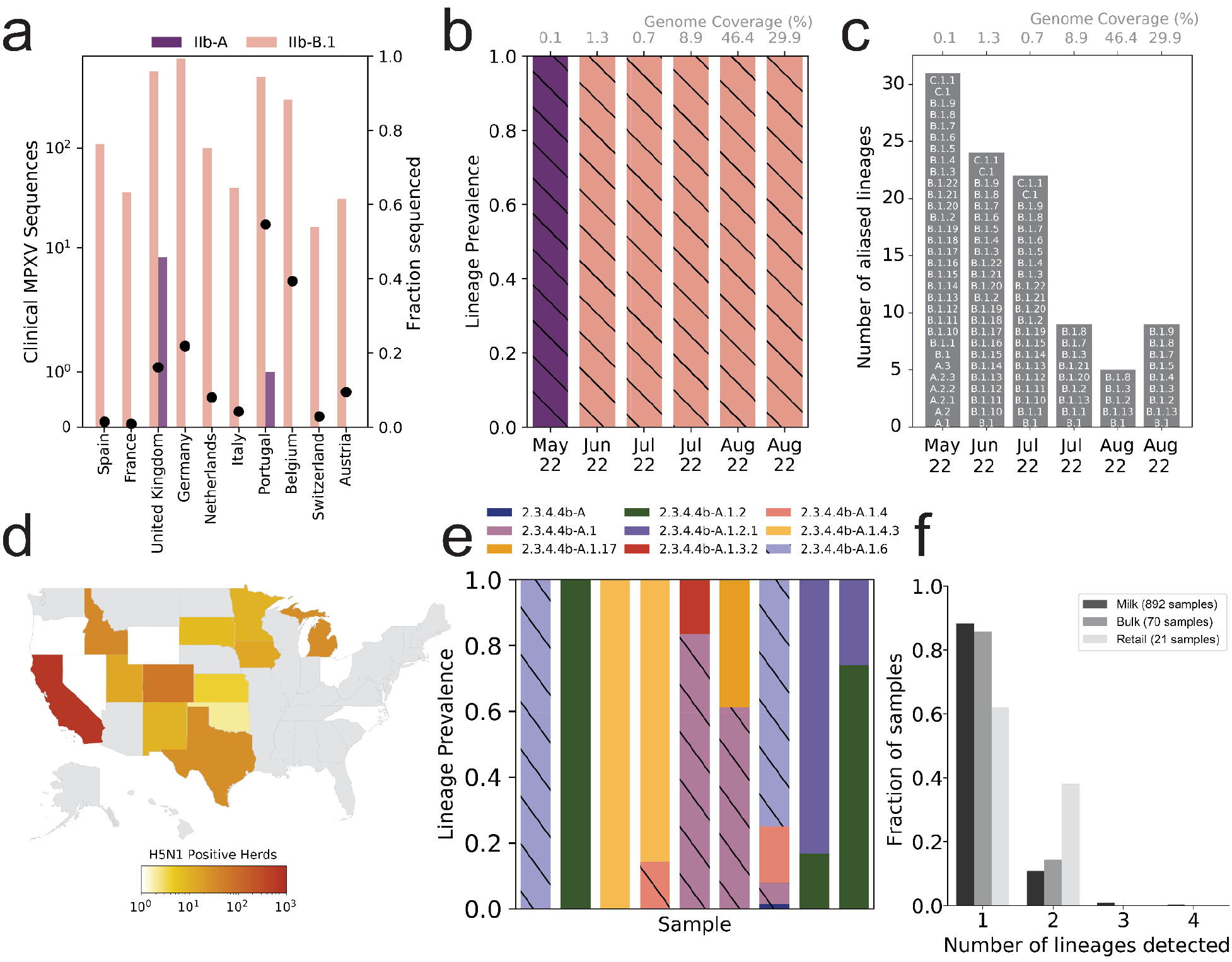
Freyja 2 enables outbreak tracking via community virus mixtures. **a**. Number of MPXV sequences by variant among European countries recording the most cases in 2022 (left axis), along with the fraction of cases for which sequencing is available (black dots, right axis). **b**. Freyja analyses identify MPXV Clade IIb-A and B.1 lineages in German wastewater, with 10x genome coverage noted above the plot. Black hatching indicates that the identified lineage corresponds to a group with the named lineage as its most recent common ancestor. **c**. The size and members of the aliased lineage groups, for each group identified in wastewater. **d**. State-level numbers of A/H5N1 positive herds identified through December 2024. **e**. Analysis of H5 2.3.4.4b lineages in milk with Freyja identifies both pure and mixed virus lineage samples. **f**. The fraction of samples with multiple distinct lineages, stratified by sample type.

## Discussion

In this study, we established Freyja 2 as a robust tool for tracking lineage prevalence and fitness of multiple pathogens directly from sequencing of wastewater and other complex samples. By accounting for key sources of error and uncertainty impacting lineage prevalence inference including sequencing errors and limited genomic coverage, Freyja 2 is stable across a wide range of sequencing platforms and approaches, providing accurate estimates of lineage prevalence while limiting false positives and reducing overall estimate error. These advances enable improved tracking of longitudinal lineage dynamics, and our lineage-specific growth rate estimation approach identifies key epidemiological information for evaluating the fitness and public health impact of emerging variants. By incorporating community surveillance resources for pathogens including MPXV and highly pathogenic avian influenza, we demonstrate that Freyja 2 can support expanded multi-pathogen wastewater and environmental surveillance, including One Health surveillance. These analyses have the potential to inform our understanding of pathogen transmission and evolution, direct epidemiological investigation, and guide public health interventions. Combined with Freyja 2’s adaptability to diverse platforms and approaches, this further supports integration of wastewater genomic surveillance of a wide range of pathogens, including in decentralized or resource-limited contexts (**Supp. Table 2)**.

Efforts to understand and enhance the potential of sequencing of wastewater and other complex samples including agricultural products and air samples for infectious disease monitoring are rapidly expanding, providing insight into pathogen dynamics that are costly or infeasible to study clinically. Recent work has leveraged wastewater for better informed phylogenetic analyses^30,67^, uncovered multiple instances of accelerated “cryptic” virus evolution^68^, and identified markers of antimicrobial resistance^69^. Lineage prevalence inference including with Freyja has been a major contributor to the expansion of wastewater genomic surveillance, and as of May 2025, Freyja has already reached over 200,000 downloads^70^.

Much like Pangolin-based lineage assignment^71^, Freyja 2 still relies on manual lineage designations from available clinical sequencing data to define pathogen lineages. Further method development is needed to enable identification of novel lineages directly from wastewater and other complex samples, including by incorporating observed read level mutation co-occurence. While development of approaches to estimate lineage effective reproduction number has the potential to further expand wastewater-based epidemiology, current knowledge of temporal pathogen shedding into wastewater is limited for all but a small subset of pathogens. As such, our proposed methods for characterization of lineage-specific growth rate are intentionally simple, though further work on understanding pathogen shedding dynamics will enable improved inference of epidemiological information.

Sequencing of many pathogens of public health interest is limited at present, especially in low-resource contexts. Genomic surveillance efforts integrating wastewater or other complex samples have the potential to substantially expand access to actionable infectious disease monitoring, and can be readily adapted to available infrastructure. As new outbreaks continue to emerge, there is a need for standardized and generalizable frameworks to analyze pathogen evolution and spread. Freyja 2’s interoperable and general architecture allows users to rapidly adapt it to new targets such as poliovirus, common cold viruses, or West Nile virus. This plug-and-play design eliminates the need to modify existing pipelines and facilitates streamlined deployment across diverse surveillance contexts. Its modular structure and open-source implementation allow easy integration into workflow managers such as Nextflow and Snakemake (**Supp. Table 2**), supporting reproducible, automated surveillance pipelines across laboratory settings. For clinical sequence data, platforms including Nextstrain allow shared phylogenetic analyses that serve as key community resources for contextualizing results. Establishing analogous tools for wastewater genomic surveillance, using methods like Freyja 2, will further increase the utility of wastewater data and provide a much-needed supplement to clinical surveillance.

## Methods

### Amplicon sequencing read simulation

All mixture simulation was performed using our Bygul simulation tool (version 1.0.0, https://github.com/andersen-lab/Bygul). Provided a list of consensus sequences and corresponding prevalences to simulate, Bygul identifies primer sites and extracts corresponding amplicons, and then uses existing read simulators including wgsim^72^ and Mason^73^ to generate the requested number of reads per sequence. For all simulation-based analyses, we generated reads with length 150 bp and set primer matching parameters to allow a maximum of one SNP mismatch per primer. Sequencing error and indel rates were set using Bygul, which supplies the appropriate parameters to the simulator. Sequence accessions CA-SDCPHL-02521150 and CA-SEARCH-201274 were used to study SNP frequency distributions, and are available on Genbank and https://github.com/andersen-lab/HCoV-19-Genomics.

### Adaptive Lasso to account for effects of sequencing error

To account for sequencing noise and counteract false positives, we modify the original formulation above to instead follow an adaptive lasso-like formulation:

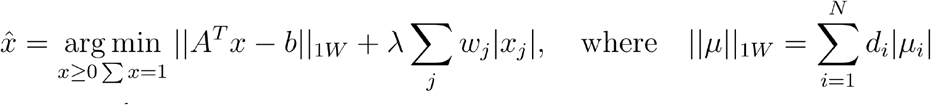

where the 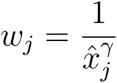 and the 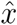 are a solution to the non-regularized problem, where we use *γ* = 1. To make the constraint term of similar scale to the residual, we first solve the unregularized problem and multiply the constraint by the unregularized residual (absorbed into lambda *λ*). This ensures that *λ* is on the same scale as the sequencing error.

### Explicit accounting of lineage identifiability due to genome coverage

To explicitly account for the level of resolution enabled by available coverage, we mask SNPs at sites without reads and grouped lineages that are no longer distinguishable from one another. That is, at genomic positions without sequencing coverage, we create the modified barcode matrix Â

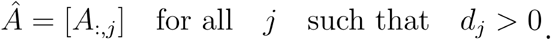

In cases where lineage rows are no longer distinguishable given the available coverage, we reduce the barcode matrix further.

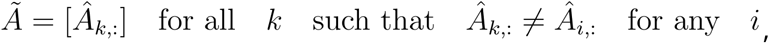

Where *k* is the representative barcode for all i such that Â_*i*,:_ = Â_*l*,:_. The resulting lineage groups are then assigned the name of their most recent common ancestor. For recombinant lineages, ancestry is traced up through the recombinant parents if groupings include lineages not from that specific recombinant.

### Freyja 2 validation and bioinformatic workflow

We selected consensus Omicron and Delta variant sequences (BCN-SEARCH-105346, CA-SEARCH-43254, CA-SEARCH-120343) collected by SEARCH researchers for validation on simulated samples (available on GenBank or https://github.com/andersen-lab/HCoV-19-Genomics). For each simulated mixture, reads were aligned to the SARS-CoV-2 reference genome (NC_045512.2) using minimap2 (v2.26-r1175) with the short-read mapping option (-ax sr), then converted to BAM files with Samtools (version 1.20). Primer sequences, low quality bases, and short reads (<50 bp) were trimmed using iVar (version 1.4.3). The processed reads were then used for variant calling using the Freyja variants command. Lineage abundances were estimated using the Freyja software alongside the SARS-CoV-2 barcode dataset (dated 2024-11-16). Freyja outputs across simulated samples were aggregated with the Freyja aggregate command, and downstream analyses were performed using custom Python and R scripts.

### Measuring robustness to sequencing error, coverage, and depth

For analyses of sequencing error or coverage, we used the wgsim simulator option in Bygul for both the three component mixture (10/30/60) as well as pairwise mixtures. The Pairwise mixture proportions ranged from 10% to 90%, and included each intermediate 10% increment for each pair of sequences. A custom Python script was used to generate amplicon dropout scenarios from 0 to 75 amplicons dropping out by subsampling the ARTIC v4.1 amplicon scheme, which includes 99 amplicons. Inference error was measured as the summed absolute value of the error between the expected and observed abundances, and sensitivity was defined as the fraction of false positives over all positives (true positives plus false negatives). Inference error was defined as the squared differences between expected and observed abundances, averaged within a 50 bp window to account for sequencing depth fluctuations. For all analyses, Freyja lineage grouping was implemented using the depthcutoff parameter (set to 1), and the autoadapt option, which uses the empirically measured error rate to set the regularization parameter.

### Lineage-specific growth rate estimation from wastewater

For a growing lineage of interest, N, with exponential growth rate *r*_*N*_ and a competing background lineage or group of lineages, B, with rate *r*_*B*_, we can model this growth as

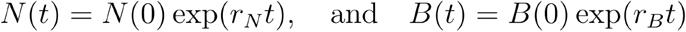

for initial abundances *N* (0) and *B* (0). Since virus nucleic acids are shed over the course of the infection, as a function of a shedding kernel *s*(*t*), the observed virus load becomes 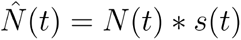. Letting the shedding kernel take the form of Gaussian, 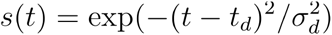, where *t*_*d*_ is the average delay in shedding following infection, and 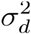 is the kernel variance. The shedding trajectory then becomes 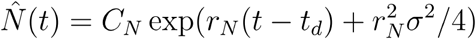 for constant *C*_*N*_ which can be simplified further to 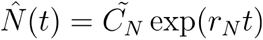.

Assuming that the shedding kernel is the same across lineages, the wastewater fraction of the lineage of interest over time can thus be written as

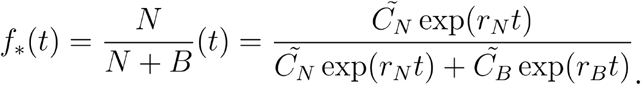

We let 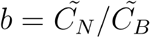 be the initial prevalence ratio of the lineage of interest relative to the background, and consider a delta distribution for generation time, *T*_*G*_ (assumed same for lineage of interest and background). We can then write the effective reproduction number *R*_*t*_ = exp (*rT*_*G*_) ^74^, and define the log growth advantage *δ* = (*r*_*N*_ − *r*_*B*_) *T*_*G*_ = log (*R*_*N*_/*R*_*B*_) such that we can rewrite the evolving wastewater lineage fraction as

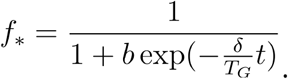

Optimizing *δ* and to maximize the logistic log-likelihood, we can then obtain the selection coefficient, *s*,

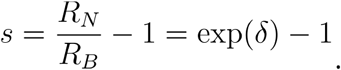

### Simulation of two-strain growth dynamics

To simulate the growth dynamics of an emerging lineage, we used a simple model of exponential phase growth in an approximately constant (i.e. S=1) susceptible population. 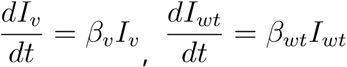. Stochastic simulation of infection dynamics was performed using a tau-leaping approach^75^, and case counts were convolved with a shedding profile obtained through LOWESS smoothing of viral loads from longitudinal fecal monitoring during COVID-19 infection^76^. To convert the lineage fraction in wastewater to read-like counts of mutation presence or absence, we used a binomial model with probability set to the fraction of the lineage of interest in the simulated wastewater signal at that time, and total number of trials set to the total number of reads occurring at a distinguishing site.

### Multi-pathogen lineage-defining mutation barcoding

For MPXV, we directly integrated the publicly available Nextstrain phylogeny and lineage designations, whereas for influenza A/H5N1 and Zika we established new nomenclature schemes. For A/H5N1, we manually identified putative lineages within the recent cattle outbreak clade, whereas for Zika we used autolin (v1.0) for automated lineage designation

^55^. To make these usable in Freyja, we converted these annotated trees into the UShER^38^ mutation annotated tree format and then used Freyja’s barcoding functionality to convert to lineage-specific barcodes. For both of these spike-in tests, we used the freyja depthcutoff parameter (set to 1), and set the adapt parameter to the approximate error rate of the sequencer (Illumina MiSeq, *λ* = 0.001; ONT R9.4.1, *λ* = 0.08). All Zika spike-in mixture sequencing used for validation was previously performed as part of previous work^77^.

### Development of a Targeted MPXV Amplicon Panel

To facilitate efficient sequencing of MPXV from wastewater, we designed a targeted amplicon panel capable of differentiating all known MPXV clades and lineages and identifying known mutations associated with Tecovirimat resistance^78^. The amplicon panel design process involved iteratively incorporating mutations defining clades or lineages, retaining only those that resulted in unique combinations for each lineage. This approach minimizes redundancy among clade-or lineage-defining mutations, while ensuring that only the most informative amplicons are included. The final panel comprises 41 amplicons and is available at https://github.com/andersen-lab/MPXV_wastewater_sequencing.

### Validation of the MPXV Amplicon Panel

We performed laboratory validation of the MPXV amplicon panel using synthetic mixtures of MPXV Clade IIb and Clade Ib. First, we determined Ct values for each clade using a pan-MPXV qPCR assay and created a series of mixtures (e.g., 90:10) of the two clades. We then sequenced technical duplicates of these mixtures using an in-house protocol ^79^. Our protocol involves an initial PCR to amplify all targeted regions, followed by a second indexing PCR. The indexing step uses the same primers as the first PCR with a universal overhang at the 5’ end, along with ONT or Illumina indexing primers that targets the universal overhang. Primer sequences, index sequences, and sequence data are all available at: https://github.com/andersen-lab/MPXV_wastewater_sequencing.

### Analysis of the spatial distribution of MPXV and Influenza A/H5N1

For MPXV, sequence data counts for each country were calculated using all available data on both GISAID and GenBank ^60^. Mpox case data for Germany and for Europe overall were downloaded directly via the WHO mpox data repository^61^. Sequencing data availability was calculated using data from For the Influenza A/H5N1 cattle outbreak, data was downloaded directly from the USDA’s HPAI Confirmed Cases in Livestock dashboard^80^.

## Code availability

Freyja is open source and available directly via github (https://github.com/andersen-lab/Freyja), bioconda (https://anaconda.org/bioconda/freyja), and as a Docker image from StaPH-B (https://hub.docker.com/r/staphb/freyja). All analyses performed in the paper are available at the project github repository (https://github.com/andersen-lab/freyja-technical-paper). BarcodeForge is available via Bioconda and GitHub (https://github.com/andersen-lab/BarcodeForge)^81^, with instructions for use available as part of the Freyja documentation (https://andersen-lab.github.io/Freyja/src/wiki/custom_barcodes.html).

## Data availability

MPXV spike-in data is available at https://github.com/andersen-lab/MPXV_wastewater_sequencing and Zika virus spike-in data can be downloaded from https://console.cloud.google.com/storage/browser/andersen-lab_project_ivar-primalseq/spike_in?invt=AbjV9g&project=andersen-lab-primary&pageState=(%22StorageObjectListTable%22:(%22f%22:%22%255B%255D%22)). Influenza A/H5N1 milk sequencing is publicly available on SRA and was obtained by the query “milk[All Fields] AND H5N1[All Fields]”. The full list of SRA Accessions used in these analyses is available in the **Supp. Data**. The MPXV sequencing data from German wastewater samples analyzed in this study are available through the NCBI Sequence Read Archive (BioProject PRJNA874069)^59^.

## Acknowledgements

We wish to thank the WHO BioHub for providing the Clade Ib MPXV isolate used for spike-in validation testing, as well as the USDA and other laboratories who have made milk sequencing data available on SRA. In particular, we’d like to thank David O’Connor and the rest of the O’Connor lab for sharing milk sequencing data and for providing helpful comments on the manuscript. We would also like to thank all of the members of Modjadji wastewater surveillance initiative and the San Diego Epidemiology and Research for COVID Health (SEARCH) alliance. This work has been funded by Centers for Disease Control and Prevention (75D30120C09795 to LCL, RK, KGA; BAA 200-2021-11554 to ASH; 75D30124C20302 to ASH), National Science Foundation (2029069 to RK), National Institutes of Health (5T32AI007244-38 to JIL; U01AI151812 to DJP; U19AI110818 to DJP; 1DP1AT010885 to RK, 3U19AI135995-03S2 to KGA; U19AI135995 to KGA; U01AI151812 to KGA, UL1TR002550 to KGA), the Gates Foundation (057213 to JIL, KGA), and the Conrad Prebys Foundation (to KGA).

## Ethics declarations

DM is a consultant for BiomeSense, Inc., has equity, and receives income. The terms of these arrangements have been reviewed and approved by the University of California, San Diego in accordance with its conflict-of-interest policies. KGA has received consulting fees for advising on SARS-CoV-2, variants and the COVID-19 pandemic. The other authors declare no competing interests.

## Supplementary Figures

**Supplementary Figure 1.**
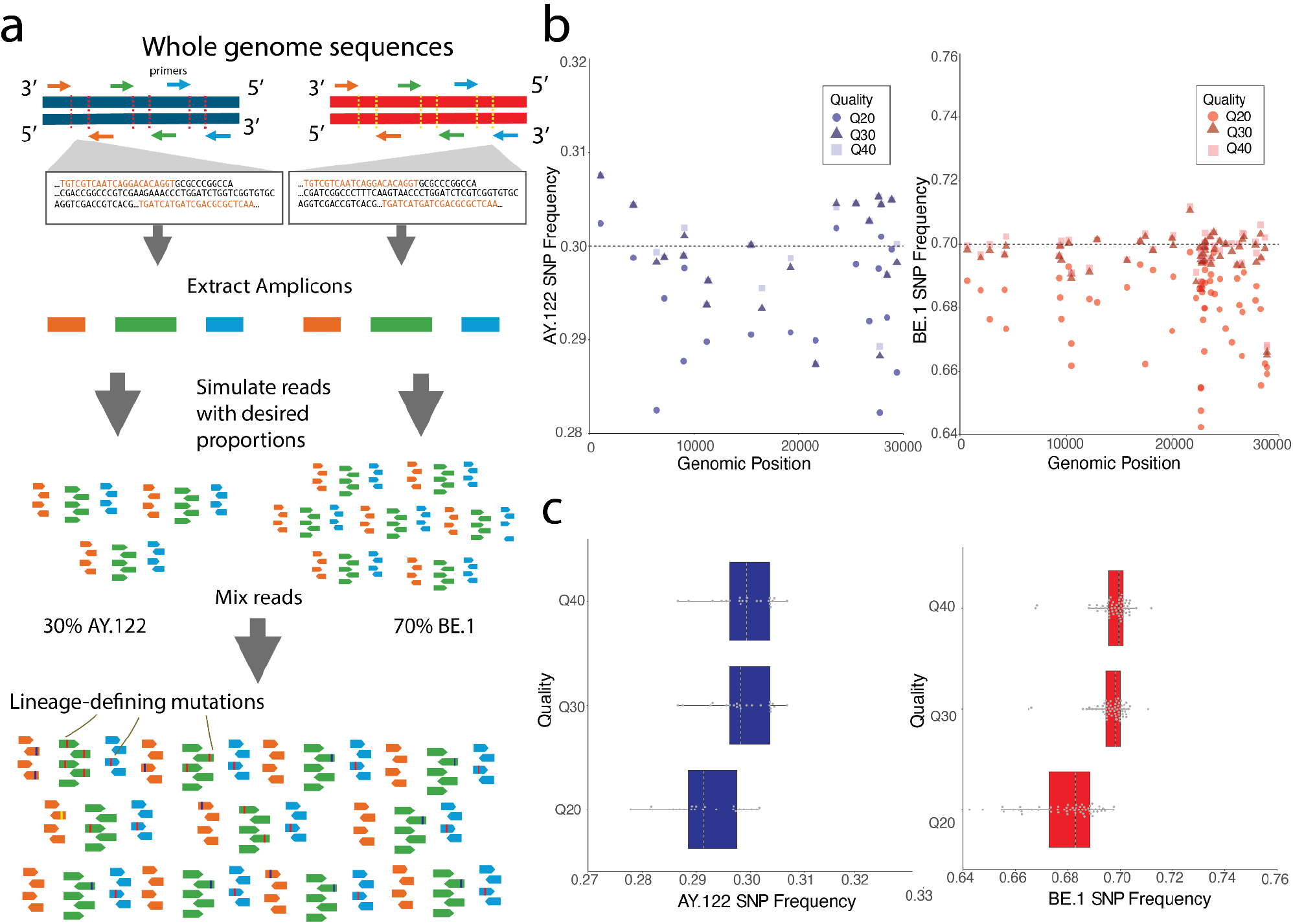
Simulating mixed lineage sequencing for Freyja 2 testing. **a**. Schematic of *in silico* amplicon sequencing simulation. **b**. Frequency of lineage-associated SNPs (defined by lineage barcodes) across the genome, for a range of sequencing qualities. Gray lines represent expected SNP frequency values. **c**. SNP frequency for repeated mixture simulations, stratified by sequencing quality. Gray dashed lines represent the median value for each boxplot, dots are the observed SNP frequency values in the mixtures.

**Supplementary Figure 2.**
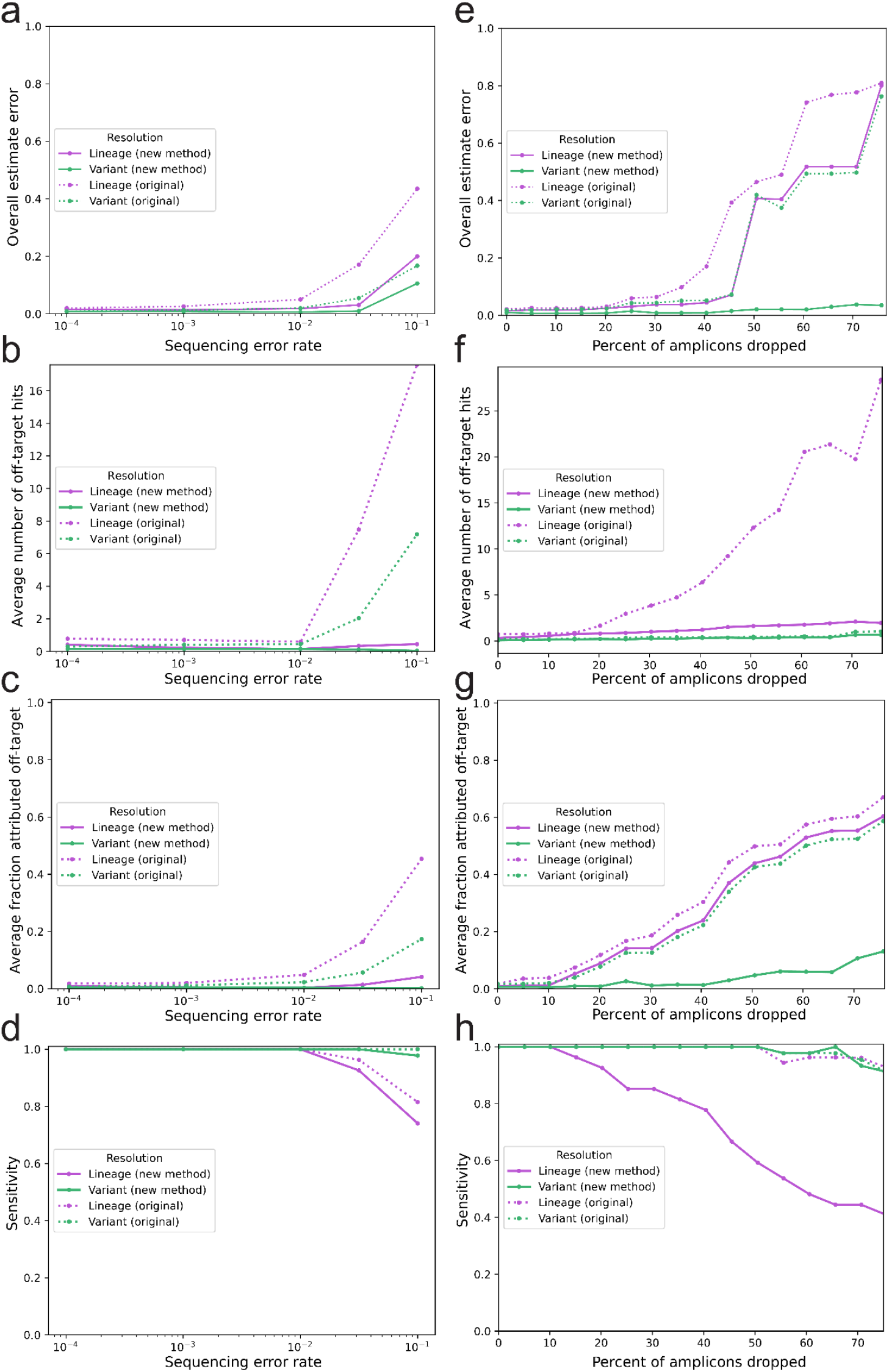
Leveraging simulation to characterize Freyja 2 A-D. Benchmarking of the Freyja 2 method against Freyja 1 (original) using simulated two-component spike-in mixtures. **a**. Estimated versus expected lineage prevalence, at varying simulated error rate. **b**. Estimated versus expected variant prevalence, aggregating identified lineages by parent (e.g. BA.2 or B.1.617.2). **c**. Inference error and **d**. sensitivity as a function of simulated sequencing error rate. **e**. Estimated versus expected lineage prevalence, with varying genomic coverage. **f**. Estimated versus expected variant prevalence, aggregating identified lineages by parent (e.g. BA.2 or B.1.617.2). **g**. Inference error and **h**. sensitivity as a function of simulated genomic coverage. **i**. Expected versus observed lineage frequencies when accounting for sequencing coverage depth. Point size and colors represent the sequencing coverage depth for each simulated mixture. **j**. Mean squared error values for each simulated pair and their respective coverage depth values. The error values are averaged using a coverage depth window size of 50 base pairs.

**Supplementary Figure 3.**
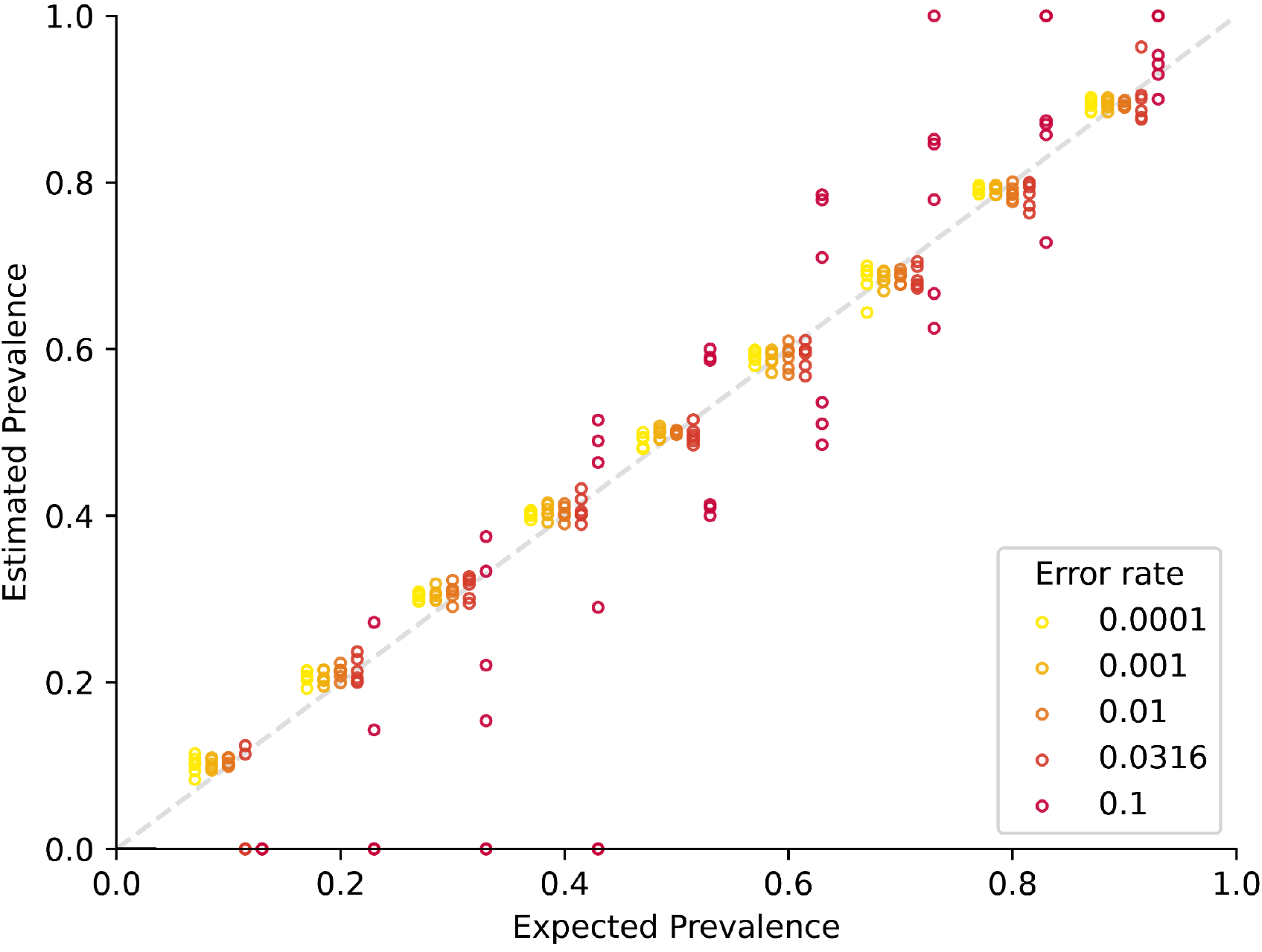
Relating sequencing error rate to estimated prevalence. Estimated vs expected (true) lineage prevalence for simulated mixtures, with values colored and by the error rate used for simulation. Samples shown are spread out horizontally by mixture type, but true mixtures prevalences are restricted to decile fractions (e.g. 10/90, 40/60).

**Supplementary Figure 4.**
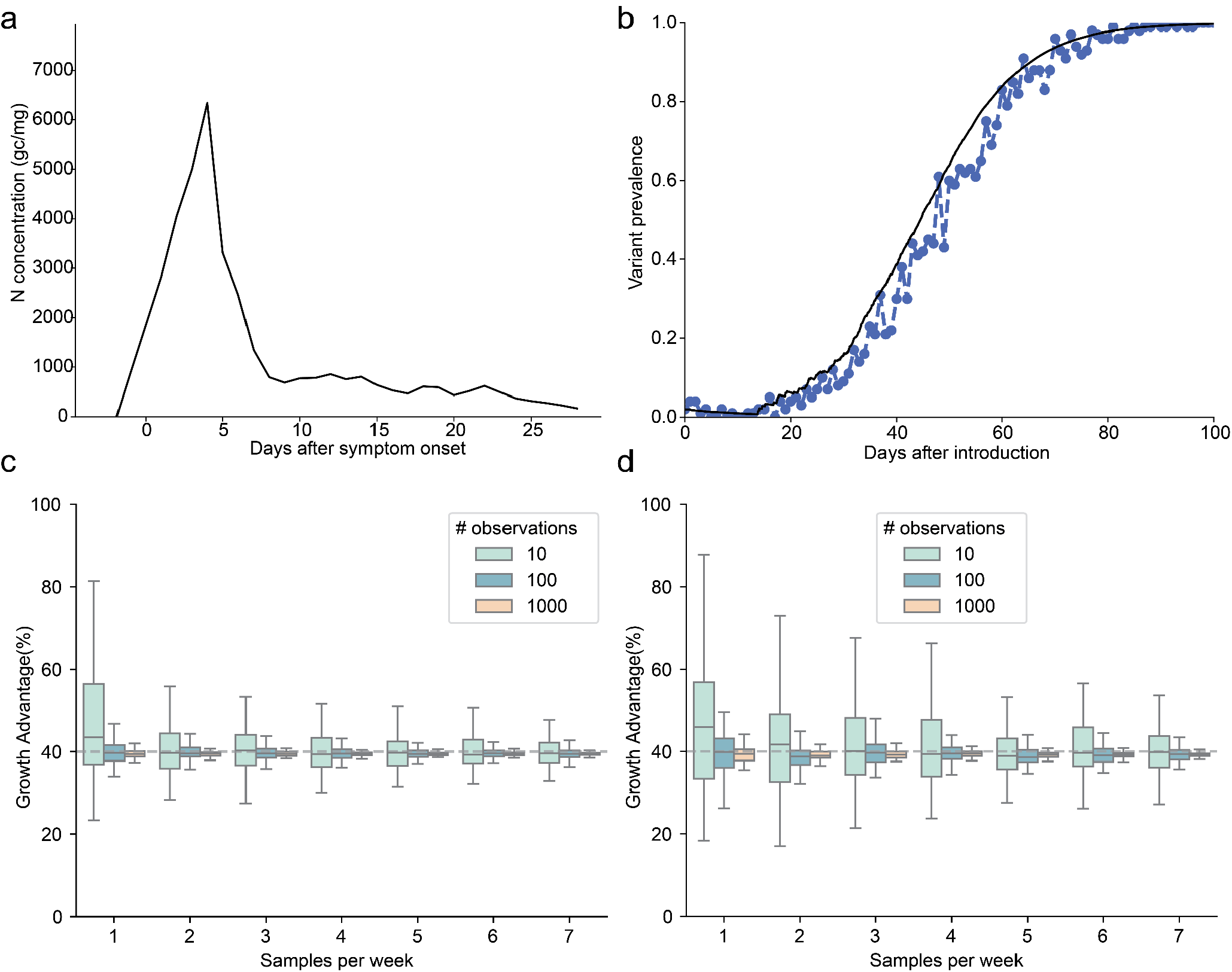
Simulation and growth rate inference emerging variant observed in wastewater. **a**. Estimated average virus shedding during SARS-CoV-2 infection as obtained from longitudinal fecal sampling^76^. **b**. Stochastic simulation of variant dynamics and corresponding wastewater signal. **c-d**. Boxplots of estimated growth advantage for the emerging variant, as a function of the number of read occurrences at a genomic site differentiating from background viruses, using the **(c)** first 100 days and **(d)** the first 50 days after introduction.

**Supplementary Figure 5.**
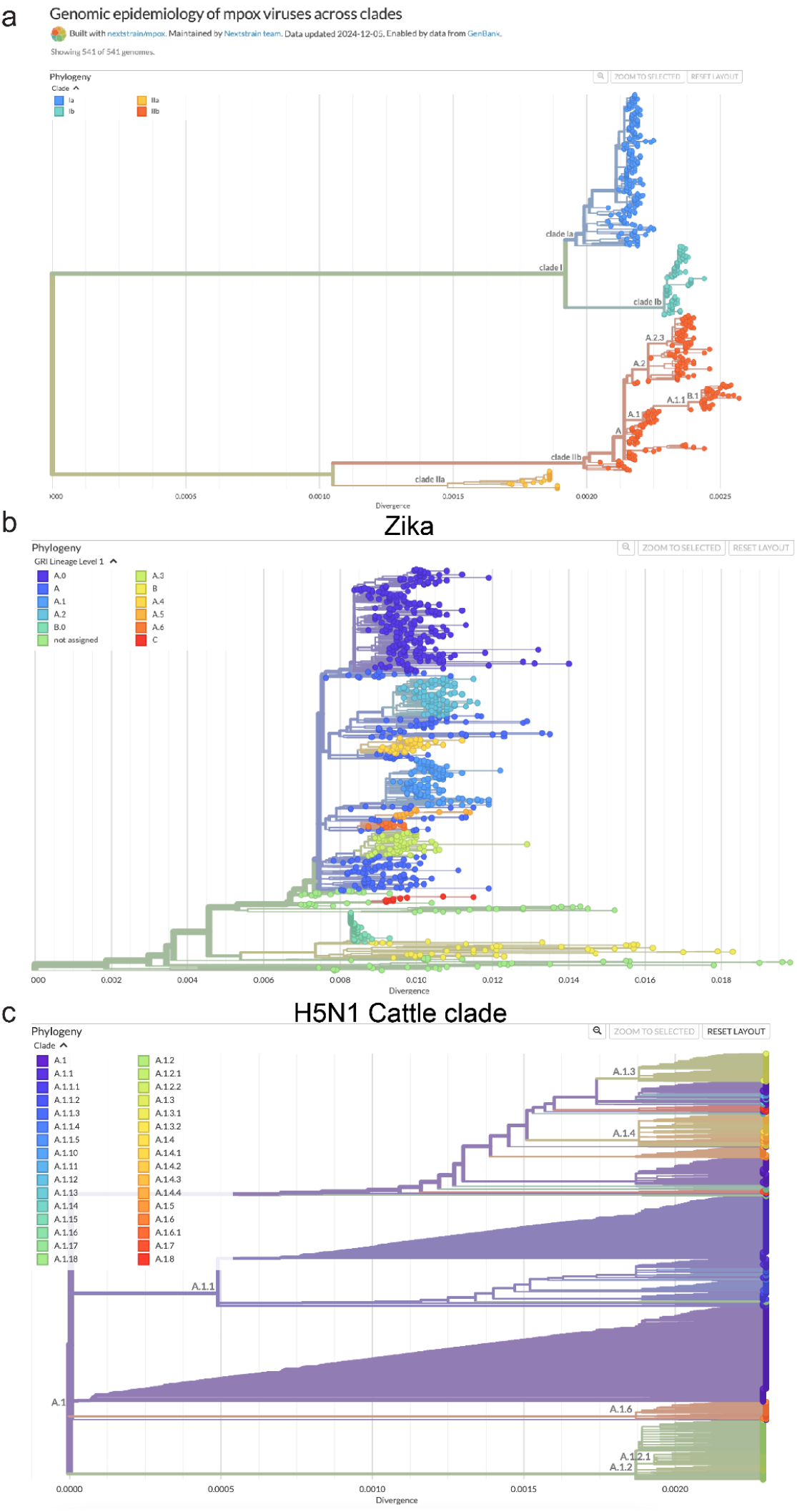
Community and custom cladistic nomenclature used for lineage barcodes. **a**. Nextstrain MPXV phylogeny with annotated clade and lineage definitions. **b**. NextStrain Zika phylogeny with ZIKV lineages defined using autolin. **c**. H5N1 cattle outbreak clade HA phylogeny with manual addition of 2.3.4.4b sub-lineages.

**Supplementary Figure 6.**
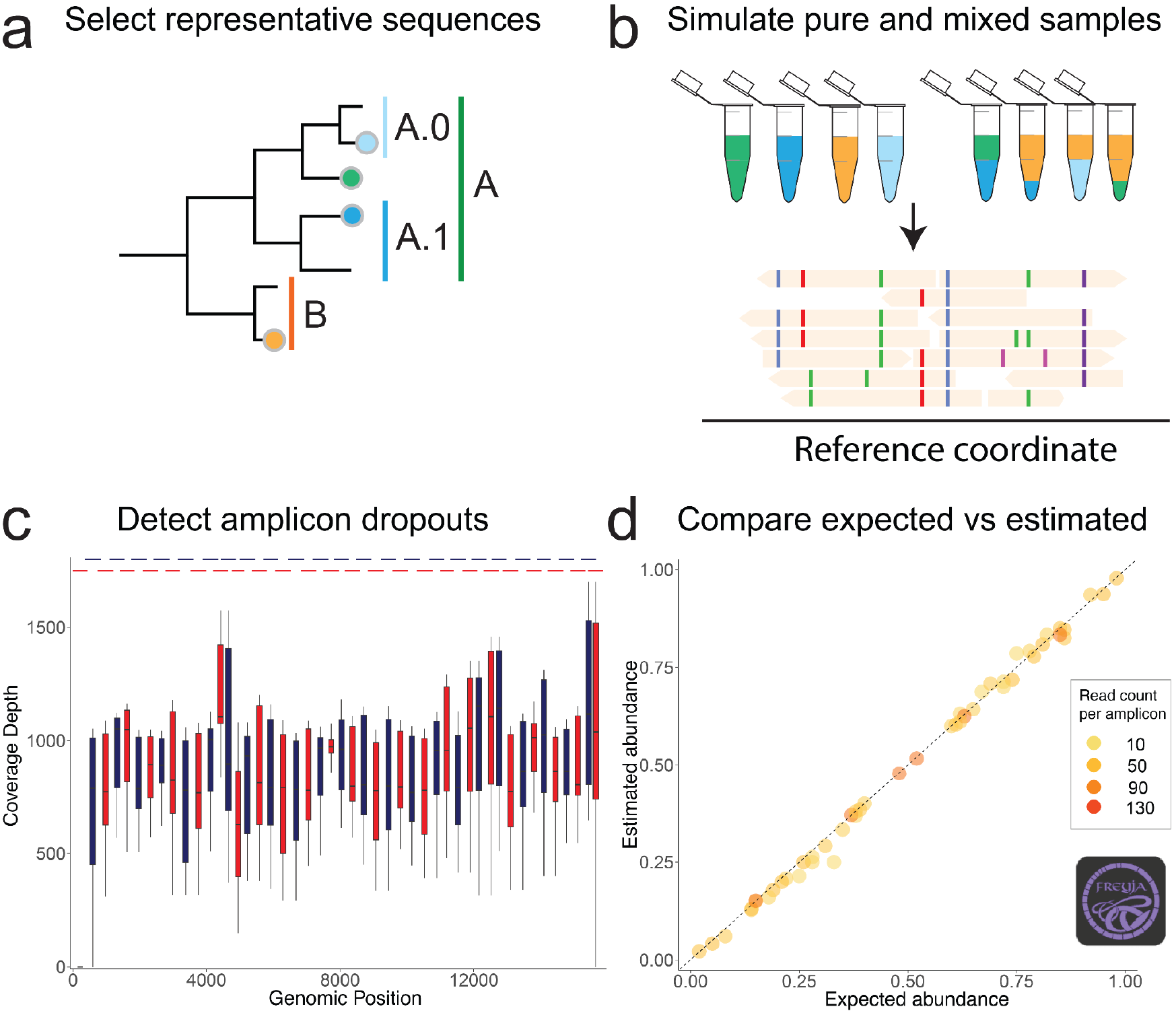
*in silico* barcode and amplicon scheme validation procedure. **a**. Representative sequences for each known clade and lineage are selected for method testing and validation. **b**. Simulation of lineage mixtures to mimic results of leading amplicon sequencing primer schemes, and mapping of reads to pathogen-specific reference. **c**. Primer scheme validation and investigation of possible lineage-specific amplicon dropouts, or likely failed amplifications, potentially resulting in missing or misleading results. **d**. Analysis of lineage mixtures with Freyja 2 to ensure expected prevalence estimates match estimates regardless of lineage sequenced.

**Supplementary Figure 7.**
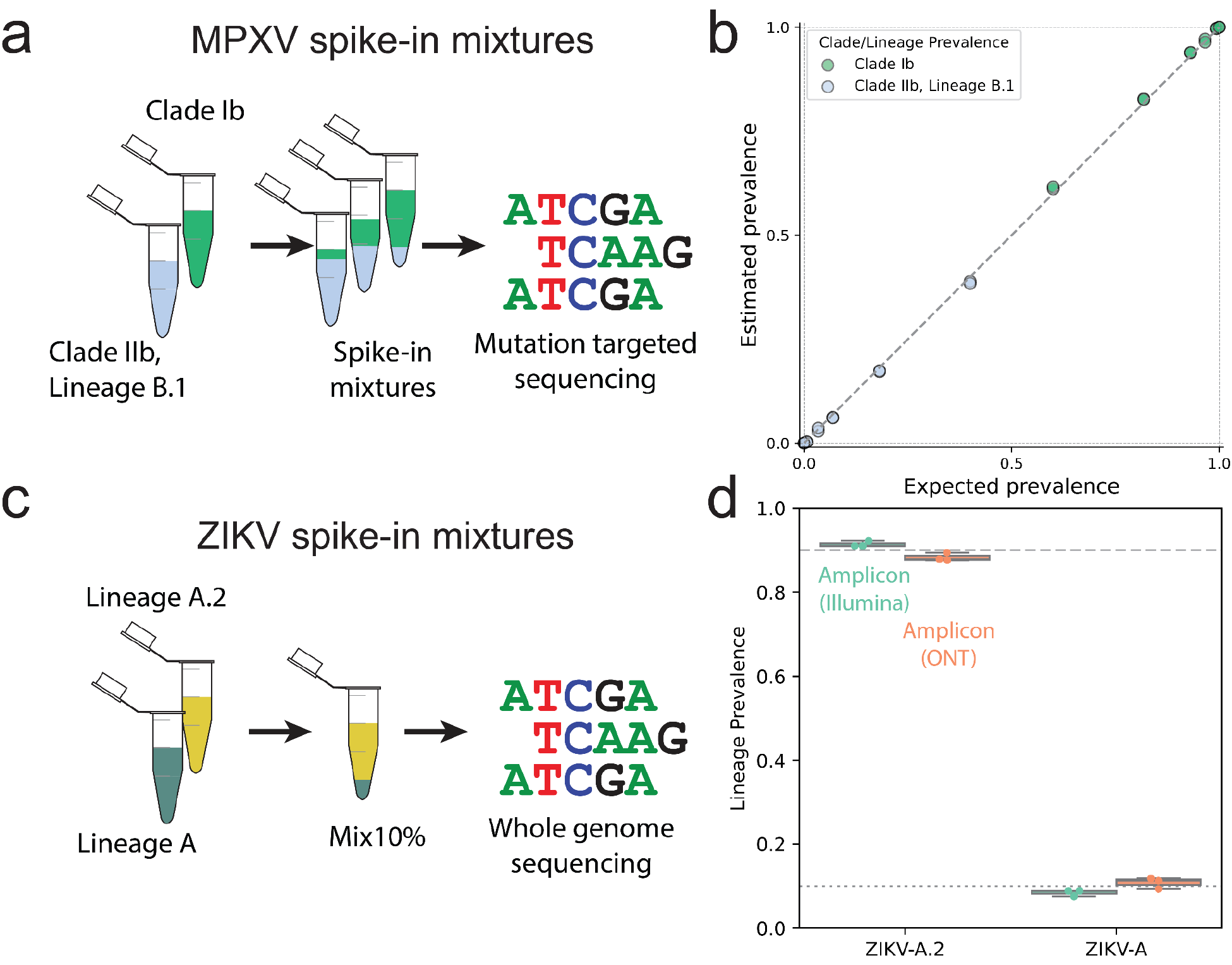
In silico and spike-in mixture laboratory validation. **a**. Schematic of the MPXV spike-in mixture experiment using Clade I and Clade IIb, Lineage B.1 virus isolates and a mutation-targeted amplicon sequencing scheme. **b**. Freyja 2 demixing on spike-in mixtures using high error rate sequencing. **c**. Schematic of the ZIKV Mix10 experiment from Grubaugh et al. 2019^77^. **d**. Deconvolution results for the Mix10 samples, for three different whole genome sequencing approaches including low error rate Illumina amplicon and high error rate Nanopore amplicon sequencing.

**Supplementary Figure 8.**
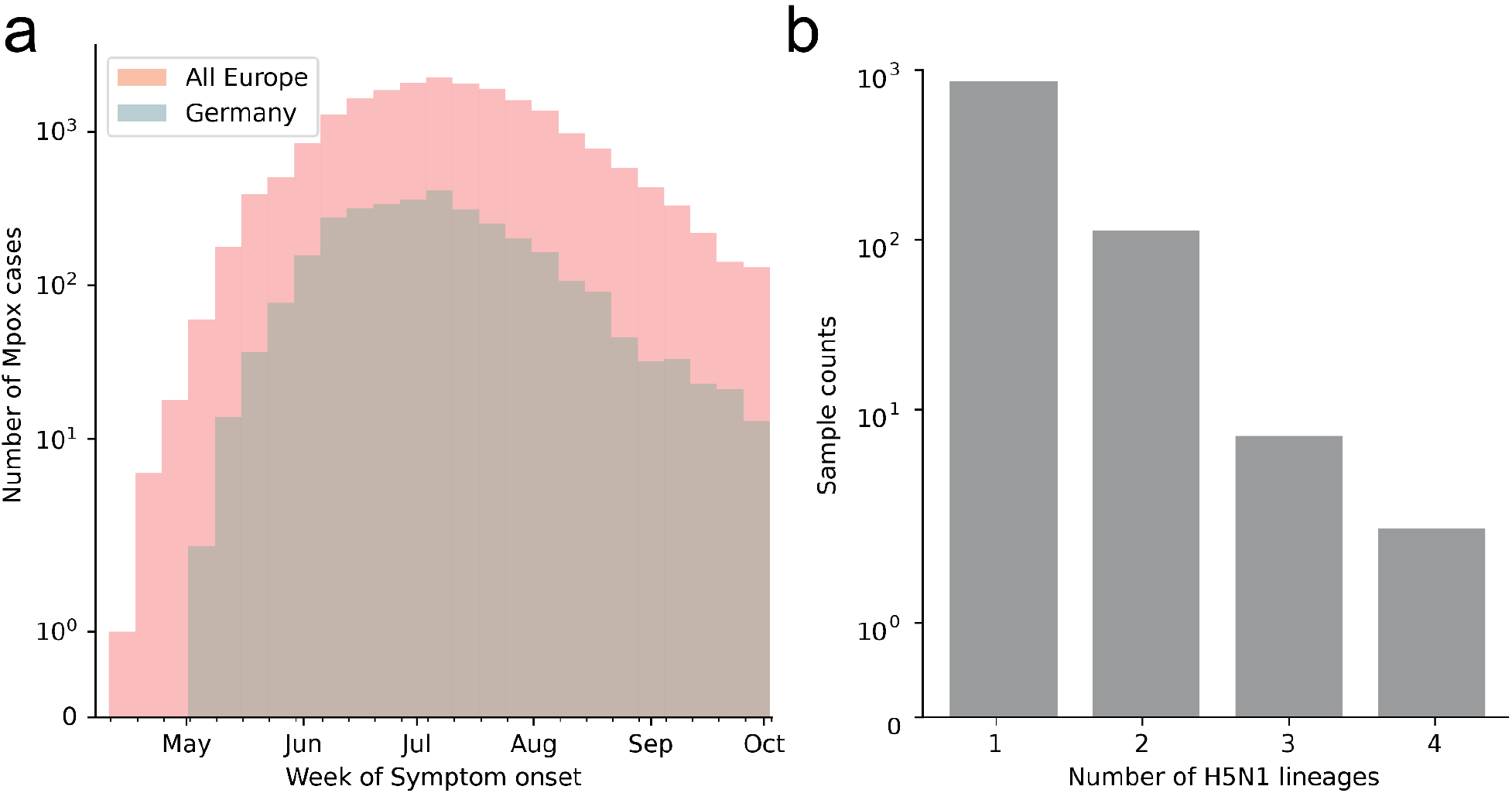
MPXV and H5N1 sequencing during outbreaks. a. Number of Mpox cases with symptom onset during the early 2022 Clade IIb outbreak in Germany and across Europe. **b**. The number of distinct lineages detected by Freyja 2 among H5N1-positive milk samples

**Supplementary Table 1.**
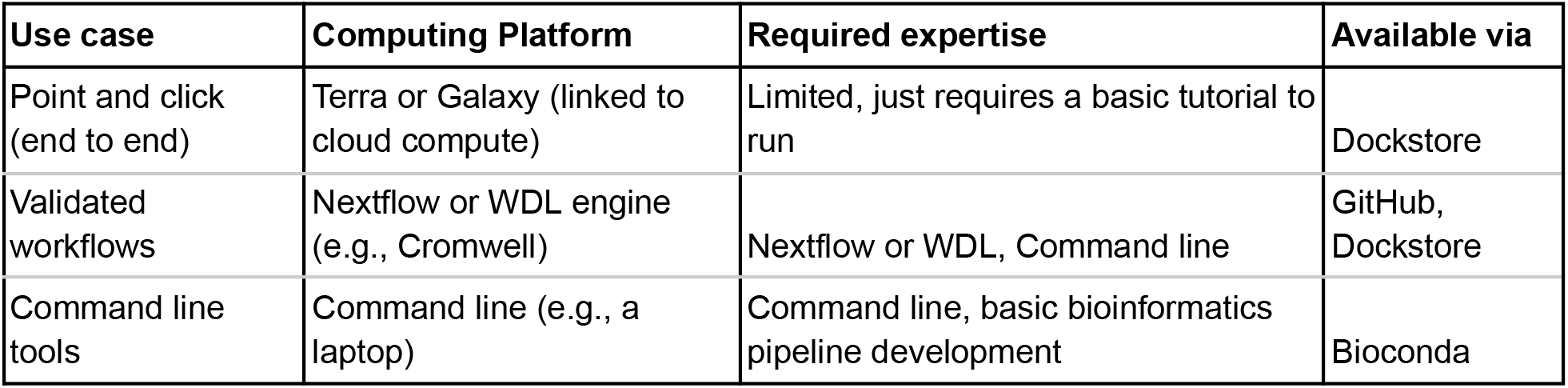
Common Freyja use cases and computing platforms.

**Supplementary Table 2.** Key differences between Freyja 1 (original) and Freyja 2. New methods and capabilities are shaded green, and improvements for enhanced accessibility, transparency, and testing are shaded yellow.

| Functionality/Properties of Method | Freyja 1 (original) | Freyja 2 |
| --- | --- | --- |
| Robustness to sequencing errors | All robustness to sequencing error derived from use of 1-norm in objective function, no regularization | Regularized objective function, with penalty scaling with empirically observed sequencing error, enabling similar results regardless of sequencing platform |
| Robustness to incomplete genome coverage | Depth-weighted inference approach, no accounting for lineage identifiability | Explicit quantification of feasible resolution of lineage inference, in addition to depth-weighted inference |
| Growth rate estimation | None | Calculation of growth rate estimation directly from Freyja analysis of longitudinal wastewater sequencing |
| Pathogen targets | SARS-CoV-2 | SARS-CoV-2, MPXV, Influenza A/H5N1, seasonal influenzas (e.g., H3N2, H1N1), RSV, measles, and more |
| Custom barcode generation workflow | Only a preset barcode generation script, exclusively for SARS-CoV-2 | BarcodeForge enables rapid generation of barcodes for other pathogens. Available as a command line tool or as a Nextflow pipeline. |
| SARS-CoV-2 Lineage barcoding | Lack of pre-processed UShER global tree, required users to download, process, and timestamp themselves | Automated daily updates by processing the UShER global tree, with paired repository for all previous barcodes, with corresponding timestamps |
| Options for Freyja access and use | Command line access via Conda or GitHub | Public health pipelines including Aquascope (Nextflow-based), point-and-click workflows via Terra, validated Docker containers via StaPH-B, and command line access via Conda and GitHub. |
| Validation tools and workflows | Laboratory spike-in mixtures | Lab spike-in mixtures, plus extensive simulation of possible mixtures with Bygul for validation |

